# The slower antibody response in myelofibrosis patients after two doses of mRNA SARS-CoV-2 vaccine calls for a third dose

**DOI:** 10.1101/2021.09.21.21263627

**Authors:** Fabio Fiorino, Anna Sicuranza, Annalisa Ciabattini, Adele Santoni, Gabiria Pastore, Martina Simoncelli, Jacopo Polvere, Sara Galimberti, Stefano Auddino, Claudia Baratè, Francesca Montagnani, Vincenzo Sammartano, Monica Bocchia, Donata Medaglini

## Abstract

Immunization with mRNA SARS-CoV-2 vaccines has been highly recommended and prioritized in fragile categories with higher risk of mortality after COVID-19 disease compared to healthy people, including patients with myelofibrosis (MF). Available data on the vaccine immune response developed by MF patients, and the impact of the treatment with the inhibitor of JAK-STAT signaling ruxolitimib, are still fragmented to support an informed decision for a third dose for this category of subjects. Here, we show that 76% of MF patients develop spike-specific IgG after the second vaccine dose, but the response has a slower kinetic compared to healthy subjects, suggesting a reduced capability of their immune system to promptly react to vaccination. A reduced ACE2/RBD inhibition binding activity of spike-specific antibodies was also observed, especially in ruxolitimib treated patients. Our results contribute to answer the open question on the induction of the antibody responses in MF patients following vaccination with COVID-19 mRNA vaccines, showing a slow kinetic that support the need for a third dose of SARS-CoV-2 mRNA vaccines.

## 1. Introduction

Onco-hematologic patients have paid a great price to COVID-19 disease caused by pandemic SARS-CoV-2 infection with a mortality rate reaching 34% of infected patients [1,2]. Immunization with mRNA SARS-CoV-2 vaccines has been highly recommended and prioritized in fragile categories including patients with hematologic malignancies (https://www.cdc.gov/coronavirus/2019-ncov/vaccines/recommendations/immuno.html), however consolidated data regarding the efficacy of anti-COVID-19 vaccines in this setting and the need of a third vaccine dose are still lacking. Patients with myelofibrosis (MF), a clonal hematopoiesis stem cells disorder belonging to the Philadelphia-negative myeloproliferative neoplasms (MPN), have a considerable higher risk of mortality after COVID-19 disease compared to the general population [1]. In addition, SARS-CoV-2 infection elicits an impaired antibody response in this subgroup of MPNs [3], similarly to other neoplastic hematologic disorders. In many MF patients the myeloproliferative disease can be controlled by the JAK1/JAK2 inhibitor ruxolitinib, a powerful inhibitor of JAK-STAT signaling that deeply reduces inflammatory cytokine production and impairs to some extent cellular immune responses [4]. Indeed, ruxolitinib has been successfully employed in dampening the cytokine storm responsible of fatal acute respiratory distress syndrome following severe COVID-19 infection, as documented by several studies [5,6]. On the other hand, the sudden interruption of ruxolitinib in MF COVID-19 infected patients has been followed by much higher death rate, probably due to the cytokine rebound following the withdraw of the drug [7].

Not enough is known about groups that might need a third vaccine dose, such as those with a compromised immune system [8]. The Centers for Disease Control and Prevention (CDC) recommends the third dose for immunocompromised subjects including blood cancer patients under treatment (https://www.cdc.gov/coronavirus/2019-ncov/vaccines/recommendations/immuno.html), while the World Health Organization (WHO) called for a moratorium on boosters until at least the end of the year (https://www.who.int/director-general/speeches/detail/who-director-general-s-opening-remarks-at-the-media-briefing-on-covid-19---8-september-2021 and https://www.who.int/news/item/10-08-2021-interim-statement-on-covid-19-vaccine-booster-doses). Several countries have or are in the process to start the third dose administration in fragile subjects despite the vaccine immune response of each category of fragile subjects has not been fully elucidated. The need and priority of a third vaccine dose in different categories of fragile patients, such as in MF patients, is a burning question, however currently available data on the vaccine response developed by MF patients are still fragmented and contradictory to support an informed decision.

Based on these premises, it is of sure interest to investigate the immune response against SARS-CoV-2 elicited by vaccination in MF patients and the potential active role of ruxolitinib in influencing this response. Up to date, preliminary data of mRNA SARS-CoV-2 vaccination observed in small groups of MPN patients are still controversial. In fact, ruxolitinib was found to interfere negatively with the generation of the humoral immune response after a single dose of mRNA vaccines in 30 MPNs patients (13 with MF) as compared to healthy subjects [9] while in another report ruxolitinib was shown not impairing the serological immune response measured after a not specified distance from second dose of BNT162b2 mRNA vaccine in 10 MF patients [10]. Different antibody responses have been reported among 42 MPN patients, with lower response to mRNA vaccine in patients with MF (10 subjects) compared to subjects affected by polycythemia vera and essential thrombocythemia [11], while another study conducted on 20 MPN subject reported that patients with a diagnosis of MF (n = 9) had significantly higher post-first dose anti-spike IgG and neutralising antibody titres compared to patients with other MPN subtypes [12].

Here, we prospectively profiled the spike-specific antibody response and the impact of the treatment with ruxolitinib in 42 consecutive MF patients, referring to our Hematology Units, at two time points (7 and 30 days) after the administration of the second dose of mRNA vaccine (Spikevax mRNA-1273 or Comirnaty BNT162b2), and in 40 healthy volunteers as control (HC).

## 2. Material and Methods

### 2.1 Study design

A cohort of 42 patients with MF, vaccinated with mRNA SARS-CoV-2 vaccine (Spikevax mRNA-1273 in 33 subjects, Comirnaty BNT162b2 in 9 subjects) and of 40 healthy vaccinated volunteers (HC) as control, was enrolled for the study. Patients with MF were treated (16/42) or not (26/42) with ruxolitinib. Plasma samples were collected at baseline (before vaccination), and 7 and 30 days from the second dose of mRNA SARS-CoV-2 vaccine received 3-4 weeks apart from the first dose. The study was performed in compliance with all relevant ethical regulations and the protocol was approved by local Ethical Committee for Clinical experimentation of Regione Toscana Area Vasta Sud Est (CEASVE), protocol code 19479 PATOVAC_COV v1.0, approved on 15 Mar 2021. All participants provided written informed consent before participation in the study. Study participants were recruited at the Hematology Unit, Azienda Ospedaliera Universitaria Senese (Siena, Italy).

### 2.2 ELISA

Maxisorp microtiter plates (Nunc, Denmark) were coated with recombinant SARS-CoV-2 full Spike protein (S1+S2 ECD, Sino Biological) with 50 μl per well of 1 μg/ml protein solution in PBS (Sigma-Aldrich) overnight at 4°C. Plates were blocked at room temperature (RT) for 1 h with 200 μl of 5% skimmed milk powder, 0.05% Tween 20, 1×PBS. All plasma samples, heated at 56°C for 1 hour to reduce risk from any potential residual virus, were added and titrated in twofold dilution in duplicate in 3% skimmed milk powder, 0.05% Tween 20, 1×PBS (diluent buffer) and incubated 1 h at RT. Anti-human horseradish peroxidase (HRP)-conjugated antibody for IgG (diluted 1:6000; Southern Biotechnology) were added in diluent buffer for 1 h at RT. Plates were developed with 3,3’,5,5’-Tetramethylbenzidine (TMB) substrate (Thermo Fisher Scientific) for 10 min at RT, followed by addition of 1M stop solution. Absorbance at 450 nm was measured on Multiskan FC Microplate Photometer (Thermo Fisher Scientific). A WHO international positive control (plasma from vaccinated donor diluted 1:5000; NIBSC) and negative control (plasma from unvaccinated donor diluted 1:20, NIBSC) were added in duplicate to each plate as internal control for assay reproducibility. Antibody end point titres were expressed as the reciprocal of the sample dilution reporting double the background OD value.

### 2.3 ACE2/RBD inhibition assay

ACE2/RBD inhibition was tested with a SARS-CoV 2 surrogate virus neutralization test (sVNT) kit (cPass™, Genscript), according to the manufacturer protocol. Briefly, plasma samples, positive and negative controls were diluted 1:10 in dilution buffer, mixed 1:1 with HRP-RBD buffer and incubated for 30 min at 37°C. 100 µl of each mixture were added to each well of a ACE2-coated 96 wells flat-bottom plate and incubated for 15 min at 37°C. The plate was washed four times with wash solution and tapped dry. 100 µl of TMB solution were added to each well and the plate was developed for 15 min at room temperature (20-25°). After that, the reaction was quenched adding 50 µl of the stop solution to each well and the OD at 450 nm was instantly read with Multiskan FC Microplate Photometer (Thermo Fisher Scientific). Results of the ACE2/RBD inhibition assay are expressed as follows: percentage inhibition = (1 - sample OD value/negative control OD value) * 100. Inhibition values ≥ 30% are regarded as positive results, while values <30% as negative results, as indicated by the manufacturer.

### 2.4 Statistical analysis

Kruskal-Wallis test, followed by Dunn’s post test for multiple comparisons, was used to assess the statistical differences of ELISA titers and ACE2/RBD inhibition percentages among at different groups of subjects for each time point post vaccination. A P-value ≤ 0.05 was considered significant. Analyses were performed using GraphPad Prism v9 (GraphPad Software, USA).

## 3. Results and Discussion

The antibody response after two doses of mRNA SARS-CoV-2 vaccine was studyed in a cohort of 42 MF patientsand 40 healthy volunteers as controls (HC). 16/42 (38%) MF patients were on ruxolitinib at the time of vaccination, while 26/42 (62%) were receiving hydroxyurea or supportive therapy only. Clinical characteristics of MF patients are outlined in Table 1.

**Table 1.** Clinical characteristics of MF patients included in the study. PV=polycythemia vera; ET= essential thrombocythemia.

| PATIENTS' CHARACTERISTICS | Whole cohort<br>(n=42) | Ruxolitinib<br>(n=16) | No Ruxolitinib<br>(n=26) |
| --- | --- | --- | --- |
| Median age at diagnosis<br>(range) | 67 years<br>(31-85 yr) | 63.5 years<br>(range 44-85 yr) | 68.5 years<br>(range 31-81 yr) |
| Sex |  |  |  |
| Male | 21 (50%) | 9 (56%) | 12 (46%) |
| Female | 21 (50%) | 7 (44%) | 14 (54%) |
| Disease |  |  |  |
| Primary MF | 21/42 (50%) | 4/21 (19%) | 17/21 (81%) |
| Post-PV | 13/42 (31%) | 9/13 (69%) | 4/13 (31%) |
| Post-ET | 8/42 (19%) | 3/8 (37.5%) | 5/8 (62.5%) |
| IPSS SCORE |  |  |  |
| LOW | 11/42 (26.2%) | 4/11 (36%) | 7/11 (64%) |
| INT-1 | 15/42 (35.8%) | 6/15 (40%) | 9/15 (60%) |
| INT-2 | 8/42 (19%) | 4/8 (50%) | 4/8 (50%) |
| HIGH | 8/42 (19%) | 4/8 (50%) | 4/8 (50%) |
| Driver mutation |  |  |  |
| JAK2 | 29/42 (69.1%) | 15/29 (51.7%) | 14/29 (48.3%) |
| CALR | 9/42 (21.4%) | 1/9 (1.1%) | 8/9 (88.9%) |
| MPL | 1/42 (2.4%) | 0/1 (0%) | 1/1 (100%) |
| Triple negative | 3/42 (7.1%) | 0/3 (0%) | 3/3 (100%) |
| Time of exposition to<br>ruxolitinib (months) |  | 26<br>(4-48 mos) |  |
| Spleen below costal margin,<br>median (range) | 3<br>(0-20 cm) | 3<br>(0-20 cm) | 3.5<br>(0-20 cm) |
| Hemoglobin g/dL, median<br>(range) | 11.8<br>(6.2-15.5) | 11.6<br>(8.5-15.5) | 12.4<br>(6.2-15.3) |
| Platelets $\times 10^3$/uL, median<br>(range) | 306<br>(19-789) | 204<br>(19-724) | 362<br>(36-789) |
| WBC $\times 10^3$/uL, median<br>(range) | 7.39<br>(2.7-37.15) | 9.47<br>(3.5-37.15) | 6.7<br>(2.7-25.27) |
| Lymphocytes $\times 10^3$/uL, median<br>(range) | 1.55<br>(0.12-7.4) | 1.49<br>(0.51-7.4) | 1.6<br>(0.12-3.03) |
| Total protein g/dL, median<br>(range) | 6.9<br>(5.8-8.5) | 7.05<br>(5.9-8.5) | 6.85<br>(5.8-7.8) |
| γ-Globulins (%), median<br>(range) | 13<br>(5.8-23.9) | 13.2<br>(9-23.9) | 12.9<br>(5.8-22) |
| LDH U/L, median<br>(range) | 415<br>(172-1430) | 377<br>(209-1019) | 498<br>(172-1430) |

Patients received the second dose of mRNA anti SARS-Cov-2 vaccine, 3-4 weeks apart from the first dose, and plasma samples were collected after 7 and 30 days (Fig. 1). Plasma samples were tested for IgG against the SARS-CoV-2 spike protein [13] by ELISA and for ACE-2/RDB inhibition activity using a SARS-CoV-2 surrogate virus neutralization test (sVNT).

**Figure 1.**
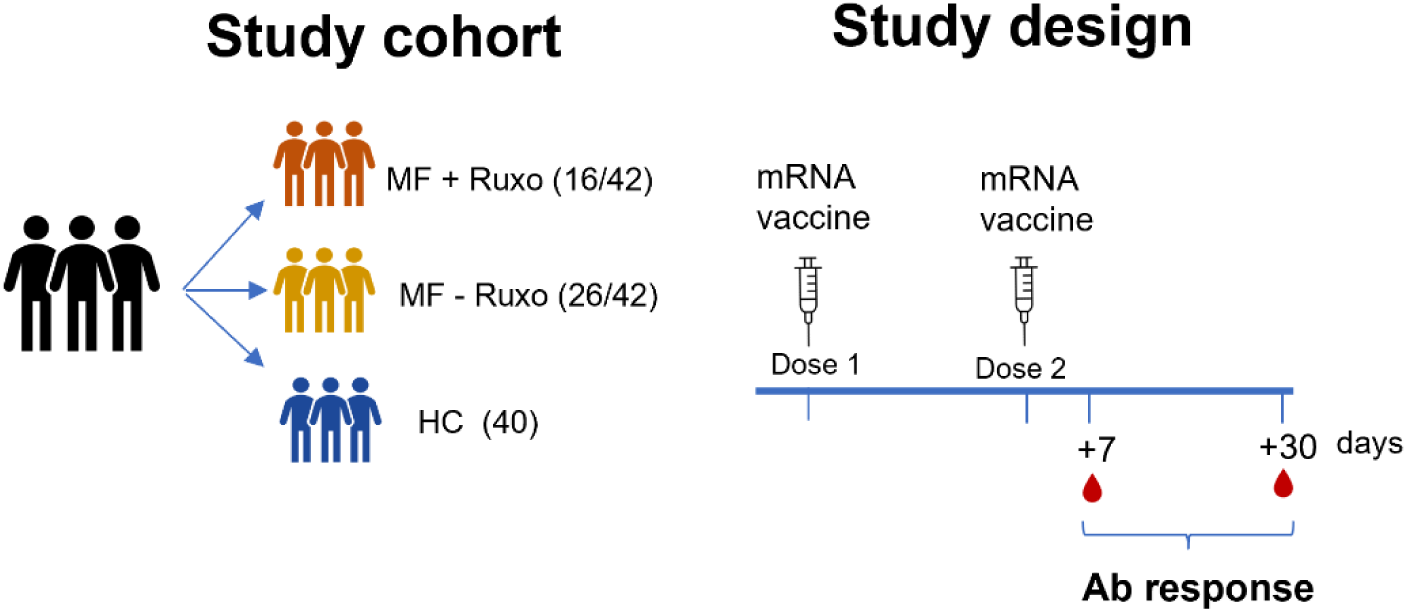
Schematic representation of the study cohort and design. Myelofibrosis (MF) patients treated or not with ruxolitinib (MF+ruxo or MF-ruxo) and Healthy controls (HC), were immunized with two doses of mRNA vaccine anti SARS-CoV-2. Antibody response was assessed 7 and 30 days after the second vaccination dose.

Seven days after the booster dose, the geometric mean tire (GMT) of anti-spike specific IgG in MF patients was significantly lower compared to HC, regardless ruxolitinib treatment (GMT of 20180, 1470 and 1060 in HC, MF-Ruxo, MF+Ruxo, respectively; P < 0.001; Figure 2A). Nevertheless, 30 days after the second dose, anti-spike IgG increased significantly in MF patients, reaching antibody titres similar to the ones observed in HC especially in the group without ruxolitinib treatment (GMT of 7896, 6267 and 2436 in HC, MF-Ruxo, MF+Ruxo, respectively; Fig. 2A-B). When MF patients were separately analyzed according to the ruxolitinib treatment, the 68% of treated and 81% of untreated patients developed a spike-specific plasma IgG response showing a slightly lower response in ruxolitinib treated subjects.

**Figure 2.**
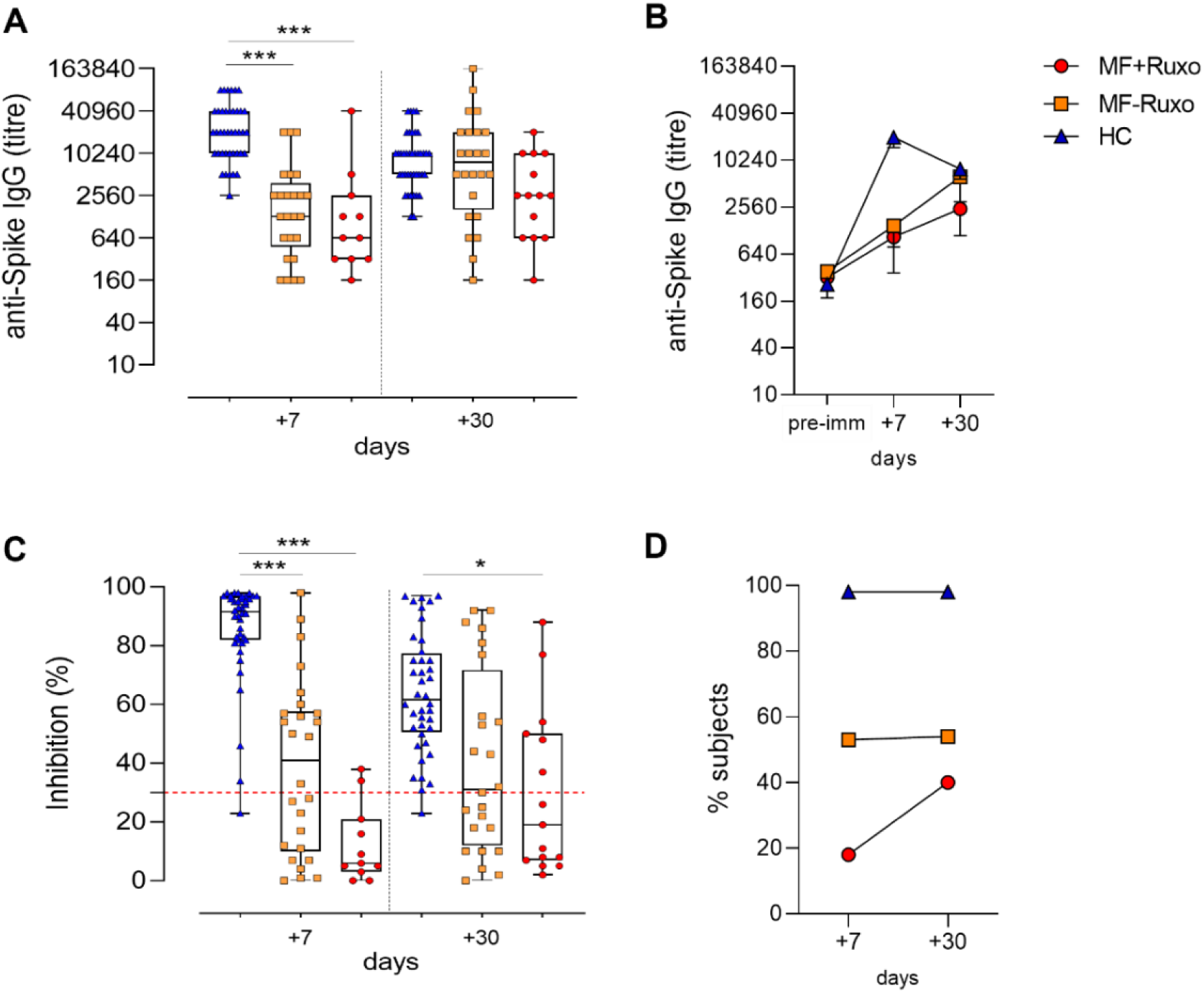
Humoral immune response against SARS-CoV-2 in MF patients post second dose of mRNA vaccination. **A)** IgG titres specific for the Spike protein assessed by ELISA in plasma of patients with myelofibrosis (MF) patients, treated or not with ruxolitinib (MF+Ruxo or MF-Ruxo), and Healthy controls (HC), at different time points post the second vaccine dose. Antibody end point titres were expressed as the reciprocal of the sample dilution reporting double the background OD value. **B)** Time course of Spike-specific IgG in plasma collected at the baseline (pre-immune) and 7 and 30 days from the second dose of mRNA SARS-CoV-2 vaccine. **C**. ACE2/RBD inhibition activity tested in plasma using a SARS-CoV-2 surrogate virus neutralization test. Results are expressed as ACE2/RBD inhibition percentage with box and whiskers diagram showing all subject values. Inhibition values >30% are regarded as positive results according to the manufacturer indication. **D**. Percentage of subjects developing ACE2/RBD inhibition positive values (>30%) in the different groups. Kruskal-Wallis test, followed by Dunn’s post test for multiple comparisons, was used for assessing statistical differences between groups. * P≤ 0.05; * * * P ≤ 0.001. Healthy control (HC), myelofibrosis (MF), ruxolitinib (Ruxo).

While the spike-specific IgG titres in HC were slightly declining a month after the booster dose (Fig. 2B), the antibodies elicited in MF patients were still increasing, thus showing a different kinetics of the secondary response, with a slower development of the humoral response in MF patients. The antibody response dynamic observed in these patients suggests a lower immunostimulating effect elicited by the first vaccine dose, as it was reported by two recent publications [9,11]. In particular, Pimpinelli et al, observed that MF subjects developed a lower humoral response compared to MNP patients with an undetectable primary immune response [11].

ACE2-RBD antibody binding inhibition, a surrogate SARS-CoV-2 neutralization test that qualitatively detects antibodies capable of inhibiting the interaction between the receptor binding domain (RBD) of the viral spike glycoprotein and the angiotensin-converting enzyme 2 (ACE2) protein [14,15], was evaluated in plasma collected 7 and 30 days after the second vaccine dose (Fig. 2C). The percentage of healthy subjects that developed a positive inhibition activity (> 30%) already 7 days after the booster dose was around 98% (39/40), and it was stably maintained at day 30 (Fig. 2D). Among MF patients, a positive inhibition activity was observed in 14/26 (54%) subjects without ruxolitinib treatment at both time points, while in ruxolitinib-treated MF patients the percentage of subjects developing antibodies with positive inhibitory activity was 18% (2/11) at day 7 and 40% (6/15) one month after the second vaccine dose (Fig. 2D)..In MF patients, expecially under ruxolitinib treatement,was therefore observed, a reduced ACE2-RBD binding inhibition activity of plasma antibodies compared to HC, even though the percentage of ruxolitinib-treated patients with a positive value increased over time. Taken together, these data demonstrate that 76% of MF patients underwent seroconversion (calculated according to WHO guidelines https://www.who.int/csr/resources/publications/WHO_CDS_CSR_ARO_2004_1.pdf) [16] (Tab. 2) after two vaccine doses, but the development of the secondary antibody response in these patients showed a slower evolution compared to HC, in contrast with the characteristics of rapidity and intensity typically associated with secondary responses to booster doses, suggesting a reduced capability of their immune system to react to vaccination. This observation is particularly relevant for defining the vaccination schedule more fitting for these patients, suggesting a possible positive role for a third vaccine dose.

**Table 2.** Number and percentage (in bracket) of patients with seroconversion and binding inhibition activity detected within MF patients (MF+Ruxo and MF-Ruxo) and HC subjects.

| Subjects | Seroconversion | Binding inhibition activity of seroconverted subjects |
| --- | --- | --- |
| Total MF | 32/42 (76%) | 22/42 (52%) |
| MF + Ruxo | 11/16 (68%) | 6/16 (37%) |
| MF - Ruxo | 21/26 (81%) | 16/26 (61%) |
| HC | 40/40 (100%) | 39/40 (98%) |

Our results, obtained in a not negligible number of MF patients prospectively studied after two doses of mRNA vaccinations, therefore shed a light on the controversial preliminary observations [9–12]. The slower antibody response observed in MF patients, regardless ruxolitinib treatment, may explain the apparent impairment of anti-Spike and neutralizing antibodies production claimed by Guglielmelli et al in MPN patients on ruxolitinib [9], and measured after only one dose of mRNA vaccines. Given our results, the JAK1/2 inhibitor treatment slightly reduces the spike-specific antibody titers elicited by two vaccine doses (Fig. 2B), but impacts on their functionality, as observed by the reduced percentage of patients with a positive ACE2-RBD binding activity (Fig. 2D). To note, none of the 42 MF patients has developed COVID-19 infection after the vaccination so far. On this regard it should be taken into consideration that MF patients “per se” experience a much higher infection rate than other MPNs patients [17] and that on the other hand, ruxolitinib seems to restore impaired monocyte functionality [18], critical for triggering the adaptive immune responses. Ongoing studies focused on the detection of Spike-specific B memory cells will further shape both the natural and vaccine–mediated anti-COVID response in this subset of fragile patients and will better clarify the grade and persistence of the immune response against SARS-CoV-2 we may expect in mRNA vaccinated MF patients.

## 4. Conclusions

Our data show that MF patients develop a humoral response after two doses of mRNA SARS-CoV-2 vaccine even if with a slower kinetics compared to healthy subjects and that a reduced antibody inhibition binding activity is observed expecially with ruxolitimib treatment. Our results therefore contribute to answer the open question on the induction of the antibody responses in MF patients following vaccination with COVID-19 mRNA vaccines and on the impact of ruxolitimib treatment. This knowledge is of critical importance to assess the need for repeated booster doses of SARS-CoV-2 mRNA vaccines and guide vaccination policies tailored for MF patients, making the case for a third vaccine dose administration.

## Data Availability

Data available upon request.

## Author Contributions

AC, DM, GP, FM, AS and MB conceived the study. GP, AS, MS, SG, CB, FM, VS and MB enrolled patients. FF, GP, JP, and SA processed the samples. AC, FF, JP carried out the immunological analysis. AC, FF, JP, SA and DM analyzed the data. AC, DM, FF and MB wrote the manuscript. AC, DM, GP and MB supervised the study. DM provided financial support. All the authors edited and approved the final version of the manuscript.

## Institutional Review Board Statement

The study was performed in compliance with all relevant ethical regulations and the protocol was approved by local Ethical Committee for Clinical experimentation of Regione Toscana Area Vasta Sud Est (CEASVE), protocol code 19479 PATOVAC COV v1.0 of 03 Mar 2021, approved on 15 Mar 2021.

## Informed Consent Statement

Informed consent was obtained from all subjects involved in the study.

## Data Availability Statement

Data available upon request.

## Acknowledgments

We would like to thank all the volunteers who participated to the study, the Ematology Unit Nursing staff who chose to cooperate for blood withdrawal. We thank Ludovica Soldateschi, Roberta Vanni and Serena Vastola for the technical support and Gianni Pozzi for critical discussion of the data.

## References

1. Passamonti, F.; Cattaneo, C.; Arcaini, L.; Bruna, R.; Cavo, M.; Merli, F.; Angelucci, E.; Krampera, M.; Cairoli, R.; Della Porta, M.G.; et al. Clinical Characteristics and Risk Factors Associated with COVID-19 Severity in Patients with Haematological Malignancies in Italy: A Retrospective, Multicentre, Cohort Study. Lancet Haematol 2020, 7, e737–e745, doi:10.1016/S2352-3026(20)30251-9.

2. Vijenthira, A.; Gong, I.Y.; Fox, T.A.; Booth, S.; Cook, G.; Fattizzo, B.; Martín-Moro, F.; Razanamahery, J.; Riches, J.C.; Zwicker, J.; et al. Outcomes of Patients with Hematologic Malignancies and COVID-19: A Systematic Review and Meta-Analysis of 3377 Patients. Blood 2020, 136, 2881–2892, doi:10.1182/blood.2020008824.

3. Passamonti, F.; Romano, A.; Salvini, M.; Merli, F.; Porta, M.G.D.; Bruna, R.; Coviello, E.; Romano, I.; Cairoli, R.; Lemoli, R.; et al. COVID-19 Elicits an Impaired Antibody Response against SARS-CoV-2 in Patients with Haematological Malignancies. British Journal of Haematology n/a, doi:10.1111/bjh.17704.

4. McLornan, D.P.; Khan, A.A.; Harrison, C.N. Immunological Consequences of JAK Inhibition: Friend or Foe? Curr Hematol Malig Rep 2015, 10, 370–379, doi:10.1007/s11899-015-0284-z.

5. Capochiani, E.; Frediani, B.; Iervasi, G.; Paolicchi, A.; Sani, S.; Roncucci, P.; Cuccaro, A.; Franchi, F.; Simonetti, F.; Carrara, D.; et al. Ruxolitinib Rapidly Reduces Acute Respiratory Distress Syndrome in COVID-19 Disease. Analysis of Data Collection From RESPIRE Protocol. Frontiers in Medicine 2020, 7, 466, doi:10.3389/fmed.2020.00466.

6. La Rosée, F.; Bremer, H.C.; Gehrke, I.; Kehr, A.; Hochhaus, A.; Birndt, S.; Fellhauer, M.; Henkes, M.; Kumle, B.; Russo, S.G.; et al. The Janus Kinase 1/2 Inhibitor Ruxolitinib in COVID-19 with Severe Systemic Hyperinflammation. Leukemia 2020, 34, 1805–1815, doi:10.1038/s41375-020-0891-0.

7. Barbui, T.; Vannucchi, A.M.; Alvarez-Larran, A.; Iurlo, A.; Masciulli, A.; Carobbio, A.; Ghirardi, A.; Ferrari, A.; Rossi, G.; Elli, E.; et al. High Mortality Rate in COVID-19 Patients with Myeloproliferative Neoplasms after Abrupt Withdrawal of Ruxolitinib. Leukemia 2021, 35, 485–493, doi:10.1038/s41375-020-01107-y.

8. Callaway, E. COVID Vaccine Boosters: The Most Important Questions. Nature 2021, 596, 178–180, doi:10.1038/d41586-021-02158-6.

9. Guglielmelli, P.; Mazzoni, A.; Maggi, L.; Kiros, S.T.; Zammarchi, L.; Pilerci, S.; Rocca, A.; Spinicci, M.; Borella, M.; Bartoloni, A.; et al. Impaired Response to First SARS-CoV-2 Dose Vaccination in Myeloproliferative Neoplasm Patients Receiving Ruxolitinib. Am J Hematol 2021, doi:10.1002/ajh.26305.

10. Caocci, G.; Mulas, O.; Mantovani, D.; Costa, A.; Galizia, A.; Barabino, L.; Greco, M.; Murru, R.; La Nasa, G. Ruxolitinib Does Not Impair Humoral Immune Response to COVID-19 Vaccination with BNT162b2 MRNA COVID-19 Vaccine in Patients with Myelofibrosis. Ann Hematol 2021, doi:10.1007/s00277-021-04613-w.

11. Pimpinelli, F.; Marchesi, F.; Piaggio, G.; Giannarelli, D.; Papa, E.; Falcucci, P.; Spadea, A.; Pontone, M.; Di Martino, S.; Laquintana, V.; et al. Lower Response to BNT162b2 Vaccine in Patients with Myelofibrosis Compared to Polycythemia Vera and Essential Thrombocythemia. J Hematol Oncol 2021, 14, 119, doi:10.1186/s13045-021-01130-1.

12. Harrington, P.; de Lavallade, H.; Doores, K.J.; O’Reilly, A.; Seow, J.; Graham, C.; Lechmere, T.; Radia, D.; Dillon, R.; Shanmugharaj, Y.; et al. Single Dose of BNT162b2 MRNA Vaccine against SARS-CoV-2 Induces High Frequency of Neutralising Antibody and Polyfunctional T-Cell Responses in Patients with Myeloproliferative Neoplasms. Leukemia 2021, 1–5, doi:10.1038/s41375-021-01300-7.

13. Ciabattini, A.; Pastore, G.; Fiorino, F.; Polvere, J.; Lucchesi, S.; Pettini, E.; Auddino, S.; Rancan, I.; Durante, M.; Miscia, M.; et al. Evidence of SARS-Cov-2-Specific Memory B Cells Six Months after Vaccination with BNT162b2 mRNA Vaccine; Frontiers in Immunology 2021; p. 2021.07.12.21259864.

14. Rincon-Arevalo, H.; Choi, M.; Stefanski, A.-L.; Halleck, F.; Weber, U.; Szelinski, F.; Jahrsdörfer, B.; Schrezenmeier, H.; Ludwig, C.; Sattler, A.; et al. Impaired Humoral Immunity to SARS-CoV-2 BNT162b2 Vaccine in Kidney Transplant Recipients and Dialysis Patients. Science Immunology 2021, 6, doi:10.1126/sciimmunol.abj1031.

15. Abe, K.T.; Li, Z.; Samson, R.; Samavarchi-Tehrani, P.; Valcourt, E.J.; Wood, H.; Budylowski, P.; Dupuis, A.P.; Girardin, R.C.; Rathod, B.; et al. A Simple Protein-Based Surrogate Neutralization Assay for SARS-CoV-2. JCI Insight 2020, 5, doi:10.1172/jci.insight.142362.

16. Long, Q.-X.; Liu, B.-Z.; Deng, H.-J.; Wu, G.-C.; Deng, K.; Chen, Y.-K.; Liao, P.; Qiu, J.-F.; Lin, Y.; Cai, X.-F.; et al. Antibody Responses to SARS-CoV-2 in Patients with COVID-19. Nat Med 2020, 26, 845–848, doi:10.1038/s41591-020-0897-1.

17. Landtblom, A.R.; Andersson, T.M.-L.; Dickman, P.W.; Smedby, K.E.; Eloranta, S.; Batyrbekova, N.; Samuelsson, J.; Björkholm, M.; Hultcrantz, M. Risk of Infections in Patients with Myeloproliferative Neoplasms—a Population-Based Cohort Study of 8363 Patients. Leukemia 2021, 35, 476–484, doi:10.1038/s41375-020-0909-7.

18. Barone, M.; Catani, L.; Ricci, F.; Romano, M.; Forte, D.; Auteri, G.; Bartoletti, D.; Ottaviani, E.; Tazzari, P.L.; Vianelli, N.; et al. The Role of Circulating Monocytes and JAK Inhibition in the Infectious-Driven Inflammatory Response of Myelofibrosis. OncoImmunology 2020.

